# Reference values for cut-point-free and traditional accelerometer metrics and associations with cardiorespiratory fitness: a cross-sectional study of healthy adults aged 20 to 89 years

**DOI:** 10.1101/2023.04.19.23288786

**Authors:** F. Schwendinger, J. Wagner, R. Knaier, D. Infanger, A.V. Rowlands, T. Hinrichs, A. Schmidt-Trucksäss

**Affiliations:** Division of Sports and Exercise Medicine, Department of Sport, Exercise and Health, University of Basel, Basel, Switzerland; Assessment of Movement Behaviours Group (AMBer), Diabetes Research Centre, Leicester General Hospital, University of Leicester, Gwendolen Rd, Leicester, UK; National Institute for Health Research (NIHR) Leicester Biomedical Research Centre (BRC), University Hospitals of Leicester NHS Trust and the University of Leicester, Leicester, UK; Alliance for Research in Exercise, Nutrition and Activity (ARENA), Sansom Institute for Health Research, Division of Health Sciences, University of South Australia, Adelaide, Australia

**Keywords:** accelerometry, physical activity, normative data, cardiorespiratory fitness, GGIR, activity monitors

## Abstract

**Objectives:** To compare the association between cardiorespiratory fitness (CRF) and cut-point-free accelerometer metrics (intensity gradient [IG] and average acceleration [AvAcc]) to that with traditional metrics in healthy adults aged 20 to 89 years and patients with heart failure, and 2) provide age-, sex-, and CRF-related reference values for healthy adults.

**Methods:** In the COmPLETE study, 463 healthy adults and 67 patients with heart failure wore GENEActiv accelerometers on their non-dominant wrist and underwent cardiopulmonary exercise testing. Cut-point-free (IG: distribution of intensity of activity across the day; AvAcc: proxy of volume of activity) and traditional (moderate-to-vigorous and vigorous activity) metrics were generated. The ‘rawacceleration’ application was developed to translate findings into clinical practice.

**Results:** IG and AvAcc yield complementary information on PA with both IG (p=0.009) and AvAcc (p<0.001) independently associated with CRF in healthy individuals. Only IG was independently associated with CRF in patients with heart failure (p=0.043). The best cut-point-free and cut-point-based model had similar predictive value for CRF in both cohorts. However, unlike traditional metrics, IG and AvAcc are comparable across populations and the most commonly used accelerometers. We produced age- and sex-specific reference values and percentile curves for IG, AvAcc, moderate-to-vigorous, and vigorous activity for healthy adults.

**Conclusions:** IG and AvAcc are strongly associated with CRF and, thus, indirectly with the risk of non-communicable diseases and mortality in healthy adults and patients with heart failure. Our reference values enhance the utility of cut-point-free metrics and facilitate their interpretation.

**Trial registration:** This study was registered on clinicaltrials.gov (NCT03986892).

- **What is already known on this topic –** Cut-point free accelerometer metrics are valuable to assess physical activity because of their comparability across populations and association with various health parameters (e.g. body fat content or physical functioning). Yet, their interpretation is not straightforward.
- **What this study adds –** This study found a strong and independent association of cut-point-free metrics with cardiorespiratory fitness, a vital sign, in healthy individuals aged between 20 to 89 years and patients with heart failure. We produced the first reference values based on healthy individuals across the age span.
- **How this study might affect research, practice or policy –** Our reference values together with the new open-source application may simplify the interpretation of cut-point-free accelerometer metrics and their use in clinical practice and research.

## INTRODUCTION

Accelerometer data can yield many physical activity (PA) outcomes.[1] Recorded acceleration is often categorised using accelerometer cut-points and expressed as minutes spent in intensity categories, i.e. light, moderate, and vigorous PA.[2] Numerous studies have proposed such cut-points for different populations.[3, 4] Although this method makes accelerometer data interpretable, the multitude of available cut-points complicates the interpretation of PA data and comparison between studies. Moreover, using cut-points to quantify the absolute intensity of PA has been shown to induce bias if cardiorespiratory fitness (CRF) of the cohort of interest differs from the cohort the cut-points were derived from.[5]

To circumvent this problem, Rowlands et al. (6) proposed cut-point-free accelerometer metrics, namely intensity gradient (IG) and average acceleration (AvAcc) and demonstrated their use across diverse populations. They moreover demonstrated the added value of using IG by examining the relation to body fatness and physical function.[6] IG was inversely associated with body fatness in adolescent girls and positively associated with physical function in adults with type 2 diabetes, independent of AvAcc.[6] The authors concluded that IG and AvAcc are appropriate for reporting as standardised measures and suitable for comparison across studies using raw-acceleration accelerometers.[6] The independent association of IG with health indicators (BMI z-scores, waist-to-height ratio, estimated CRF, and metabolic syndrome score) was later confirmed in children.[7]

CRF, measured as peak oxygen uptake (VO_2peak_), is a strong, independent risk predictor for various non-communicable diseases and all-cause mortality.[8] Data on the relationship between cut-point-free accelerometer metrics and CRF in healthy adults or diseased populations e.g. patients with heart failure would therefore be relevant. Moreover, IG and AvAcc are not readily interpretable.[6] Reference values providing a framework for judging the appropriateness of PA levels are required.[6]

Thus, we aimed to 1) examine the relationship between cut-point-free and traditional accelerometer metrics, 2) compare the association between VO_2peak_ and cut-point-free metrics to that with traditional metrics in healthy individuals and patients with heart failure, and 3) produce reference values for cut-point-free and traditional accelerometer parameters for healthy adults.

## METHODS

### Study design

Data were collected in the population-based, cross-sectional COmPLETE study at the Department of Sport, Exercise and Health at the University of Basel, Switzerland. The study protocol is available elsewhere.[9] All procedures were performed according to the Declaration of Helsinki. The study was registered on clinicaltrials.gov (NCT03986892) and approved by the Ethics Committee of North-western and Central Switzerland (EKNZ 2017– 01451).

### Study participants

The COmPLETE study examined healthy adults (HEALTHY) and patients with chronic heart failure (HEART). HEALTHY were ≥20 years of age, non-smoking, and had a BMI <30 kg/m^2^.[9] Exclusion criteria were the presence of chronic exercise-limiting disease, known pregnancy or breastfeeding, hypertonic blood pressure >160/100 mmHg, orthopaedic problems hindering the examinations, inability to follow the study procedures, and contraindications for all-out exercise.[9] Individuals aged ≥20 years and diagnosed with stable chronic heart failure were eligible for HEART.[9] All exclusion criteria and a rationale for the study size are presented elsewhere.[9]

### Study procedures

#### Accelerometry and data processing

Participants were asked to wear a triaxial accelerometer (GENEActiv, Activinsights Ltd., Kimbolton, UK) on their non-dominant wrist, for 24 hours/day over 14 days.[9] Data were sampled at 50 Hz. Days with wear time >14 hours were considered valid. Participants needed to have at least four valid weekdays (Monday to Friday) and one Saturday and Sunday each.[5] Raw-data processing using the GGIR package[10, 11] is described in detail in the Supplement.

#### Cardiopulmonary exercise testing

Gas exchange was measured breath-by-breath during a graded exercise test on a cycle ergometer using the MetaMax 3B portable metabolic system (Cortex Biophysik GmbH, Leipzig, Germany). One of five ramp protocols was chosen depending on the participant’s estimated CRF.[9] VO_2peak_ was recorded as the three highest consecutive values (30 s average) during the test. Detailed information are presented elsewhere.[12]

### Statistical analyses

Statistical analyses and figures were done in R version 4.2.0.[13] Correlations between accelerometer parameters and VO_2peak_ were analysed using Spearman’s Rank Correlation. Explorative principal component and partial least squares analyses were performed to examine the relationship between the abovementioned variables.[14, 15] Multiple linear regression models were used to examine between-cohort differences and the association between VO_2peak_ and accelerometer parameters, i.e. IG, AvAcc, MVPA, VPA and time in PA (TPA=light PA+MVPA). For each cohort, we planned to create three models with cut-point-free metrics only (IG, AvAcc, and both parameters together), two models with cut-point-based metrics only (MVPA and VPA), and two models containing IG or VPA as a measure of PA intensity and TPA as a surrogate of time in PA. All models were adjusted for age and an interaction between BMI and sex. We used restricted cubic splines with four knots placed at appropriate percentiles of the data for age and BMI.[16] Fulfilment of model assumptions was verified using residual diagnostics. The best model fit was selected based on residual standard deviation and adjusted R^2^. A Likelihood-Ratio-Test was used to compare models. Non-nested models were compared using the Vuong Test.[17] Percentile curves were created with Generalised Additive Models for Location, Scale, and Shape with the GAMLSS package (version 5.3-4).[18] Age or VO_2peak_ as the independent parameter was fitted using p-splines.[19, 20] We adopted the Bayesian information criterion to select the conditional distribution offering the best compromise between model complexity and goodness-of-fit. Model fits were inspected using diagnostic residual plots.[21] We developed the application ‘rawacceleration’ using ‘shiny’[22] to translate our findings into clinical practice. The level of statistical significance was set at 0.05. All tests were two-sided.

## RESULTS

679 HEALTHY and 89 HEART were invited to this study. Of those, 585 HEALTHY and 70 HEART fulfilled all criteria. After excluding participants with missing covariate data, 463 HEALTHY and 67 HEART were analysed. Cohort characteristics are presented in Table 1.

**Table 1.**
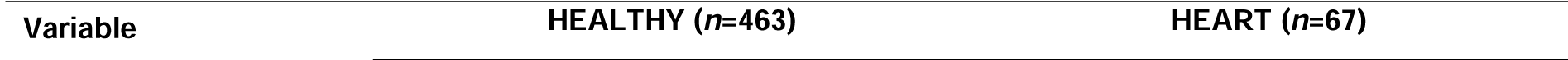
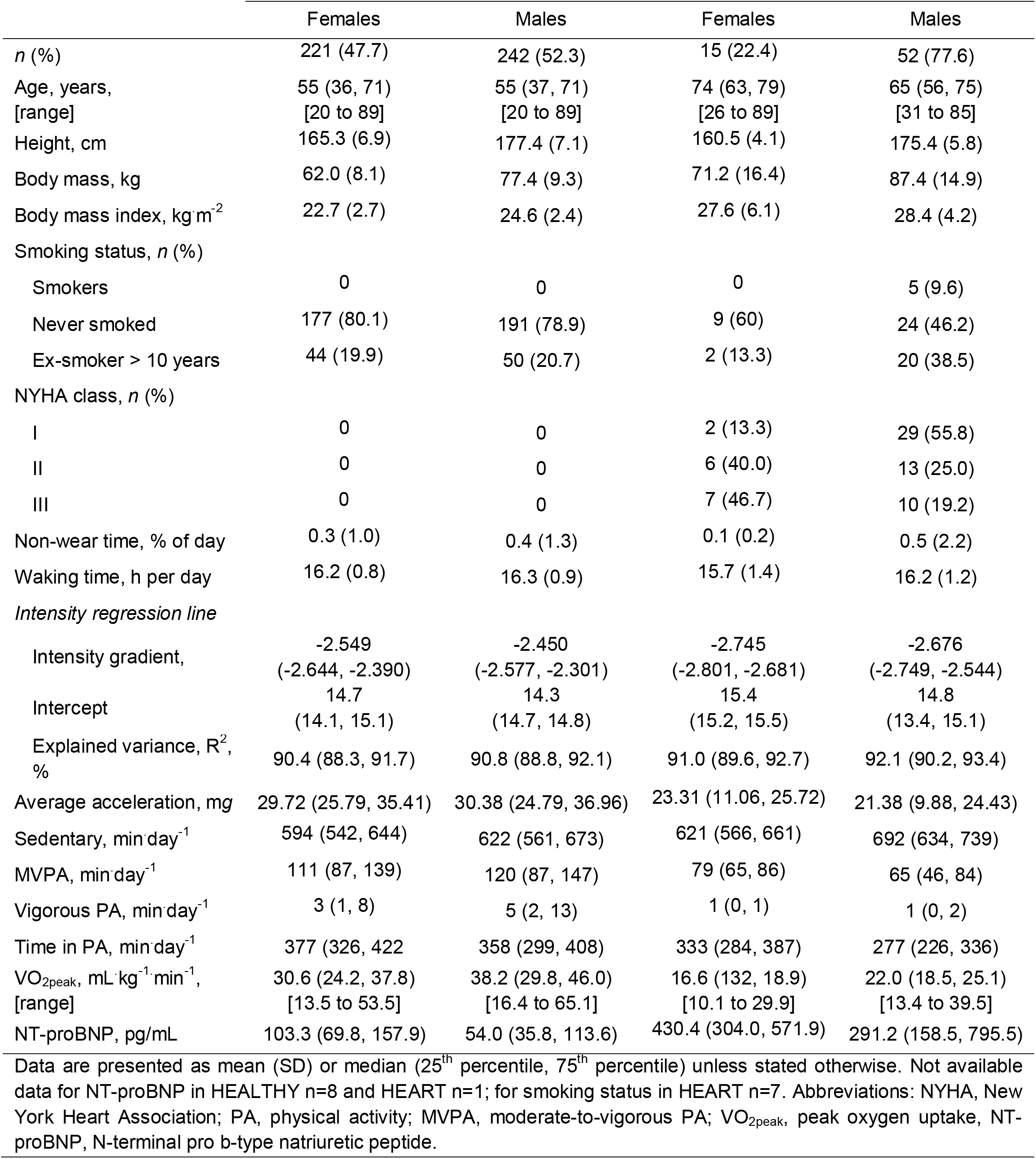
Characteristics of healthy individuals and patients with heart failure stratified by sex.

|  |  |  |
| --- | --- | --- |
| Variable | HEALTHY (n=463) | HEART (n=67) |

|  | Females | Males | Females | Males |
| --- | --- | --- | --- | --- |
| *n* (%) | 221 (47.7) | 242 (52.3) | 15 (22.4) | 52 (77.6) |
| Age, years,<br>[range] | 55 (36, 71)<br>[20 to 89] | 55 (37, 71)<br>[20 to 89] | 74 (63, 79)<br>[26 to 89] | 65 (56, 75)<br>[31 to 85] |
| Height, cm | 165.3 (6.9) | 177.4 (7.1) | 160.5 (4.1) | 175.4 (5.8) |
| Body mass, kg | 62.0 (8.1) | 77.4 (9.3) | 71.2 (16.4) | 87.4 (14.9) |
| Body mass index, kg·m<sup>-2</sup> | 22.7 (2.7) | 24.6 (2.4) | 27.6 (6.1) | 28.4 (4.2) |
| Smoking status, *n* (%) |  |  |  |  |
| Smokers | 0 | 0 | 0 | 5 (9.6) |
| Never smoked | 177 (80.1) | 191 (78.9) | 9 (60) | 24 (46.2) |
| Ex-smoker > 10 years | 44 (19.9) | 50 (20.7) | 2 (13.3) | 20 (38.5) |
| NYHA class, *n* (%) |  |  |  |  |
| I | 0 | 0 | 2 (13.3) | 29 (55.8) |
| II | 0 | 0 | 6 (40.0) | 13 (25.0) |
| III | 0 | 0 | 7 (46.7) | 10 (19.2) |
| Non-wear time, % of day | 0.3 (1.0) | 0.4 (1.3) | 0.1 (0.2) | 0.5 (2.2) |
| Waking time, h per day | 16.2 (0.8) | 16.3 (0.9) | 15.7 (1.4) | 16.2 (1.2) |
| *Intensity regression line* |  |  |  |  |
| Intensity gradient, | -2.549<br>(-2.644, -2.390) | -2.450<br>(-2.577, -2.301) | -2.745<br>(-2.801, -2.681) | -2.676<br>(-2.749, -2.544) |
| Intercept | 14.7<br>(14.1, 15.1) | 14.3<br>(14.7, 14.8) | 15.4<br>(15.2, 15.5) | 14.8<br>(13.4, 15.1) |
| Explained variance, R<sup>2</sup>,<br>% | 90.4 (88.3, 91.7) | 90.8 (88.8, 92.1) | 91.0 (89.6, 92.7) | 92.1 (90.2, 93.4) |
| Average acceleration, mg | 29.72 (25.79, 35.41) | 30.38 (24.79, 36.96) | 23.31 (11.06, 25.72) | 21.38 (9.88, 24.43) |
| Sedentary, min·day<sup>-1</sup> | 594 (542, 644) | 622 (561, 673) | 621 (566, 661) | 692 (634, 739) |
| MVPA, min·day<sup>-1</sup> | 111 (87, 139) | 120 (87, 147) | 79 (65, 86) | 65 (46, 84) |
| Vigorous PA, min·day<sup>-1</sup> | 3 (1, 8) | 5 (2, 13) | 1 (0, 1) | 1 (0, 2) |
| Time in PA, min·day<sup>-1</sup> | 377 (326, 422) | 358 (299, 408) | 333 (284, 387) | 277 (226, 336) |
| VO<sub>2peak</sub>, mL·kg<sup>-1</sup>·min<sup>-1</sup>,<br>[range] | 30.6 (24.2, 37.8)<br>[13.5 to 53.5] | 38.2 (29.8, 46.0)<br>[16.4 to 65.1] | 16.6 (132, 18.9)<br>[10.1 to 29.9] | 22.0 (18.5, 25.1)<br>[13.4 to 39.5] |
| NT-proBNP, pg/mL | 103.3 (69.8, 157.9) | 54.0 (35.8, 113.6) | 430.4 (304.0, 571.9) | 291.2 (158.5, 795.5) |

Data are presented as mean (SD) or median (25<sup>th</sup> percentile, 75<sup>th</sup> percentile) unless stated otherwise. Not available data for NT-proBNP in HEALTHY n=8 and HEART n=1; for smoking status in HEART n=7. Abbreviations: NYHA, New York Heart Association; PA, physical activity; MVPA, moderate-to-vigorous PA; VO<sub>2peak</sub>, peak oxygen uptake, NT-proBNP, N-terminal pro b-type natriuretic peptide.

### Between-cohort differences

HEART had significantly lower IG (p<0.001, 95% CI: -0.185 to -0.079), AvAcc (p<0.001; 95% CI: -8.59 to -3.92 m*g*), MVPA (p<0.001; 95% CI: -45 to -20 min), VPA (p=0.002; 95% CI: -6.1 to -1.3 min), and TPA (p<0.001; 95% CI: -80 to -33 min) than HEALTHY (see Table 1). HEART were more sedentary than HEALTHY (p=0.006; 95% CI: 10 to 59 min).

### Interrelationship between accelerometer metrics and VO_2peak_ in HEALTHY and HEART

Correlations of VO_2peak_, IG, AvAcc, MVPA, VPA, TPA, and sedentary time are available in Figure 1.

**Figure 1.**
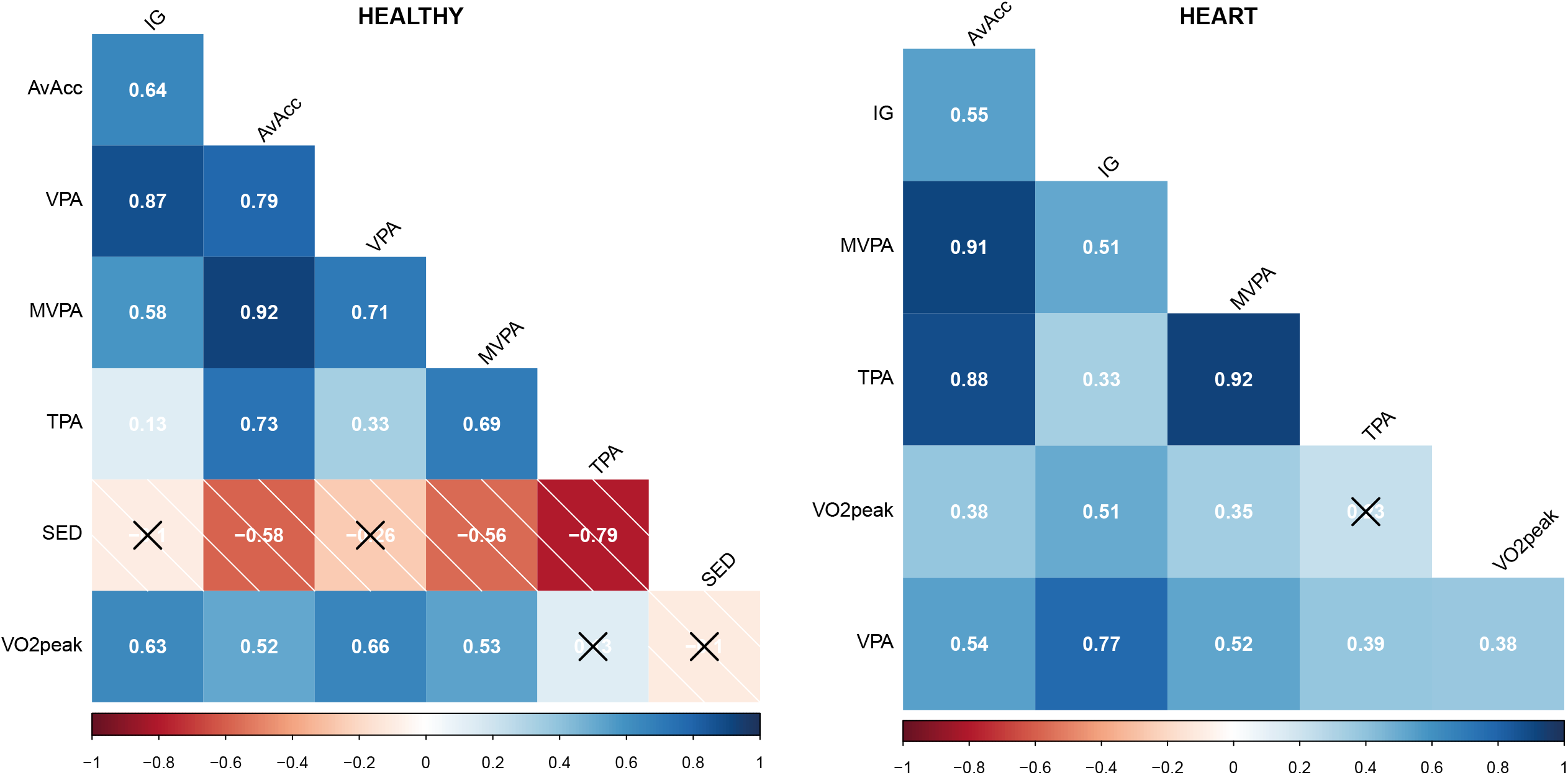
Spearman correlations of average acceleration (AvAcc), intensity gradient (IG), moderate-to-vigorous physical activity (MVPA), sedentary time (SED), time in physical activity (TPA), peak oxygen uptake (VO2peak), and vigorous physical activity (VPA) for healthy individuals (HEALTHY) and patients with heart failure (HEART), respectively. Non-significant correlations are crossed.

Results of the explorative principal component and partial least squares analyses are presented and discussed in Supplement 2.

Multiple regression models with key accelerometer parameters as independent variables and VO_2peak_ as the dependent variable are presented in Table 2. In HEALTHY, model 2 (AvAcc) had a better fit than model 1 (IG) (p=0.022). Model 7 (AvAcc+IG) performed better than model 1 (IG) (p<0.001) and model 2 (AvAcc) (p=0.004). The best cut-point-free model including AvAcc and IG (model 7) and the best cut-point-based model including VPA (model 4) performed similarly.

**Table 2.** Regression models for HEALTHY and HEART with peak oxygen uptake as the dependent variable.

| Independent variables | Estimate | 95% CI | p-value | Residual standard deviation | Adjusted R<sup>2</sup> |
| --- | --- | --- | --- | --- | --- |
| *HEALTHY* (n=463) |  |  |  |  |  |
| <u>Model 1</u> |  |  | <0.001 | 5.47 | 0.7153 |
| IG | 10.8 | (7.9 to 13.8) | <0.001 |  |  |
| <u>Model 2</u> |  |  | <0.001 | 5.27 | 0.7361 |
| AvAcc | 0.3 | (0.24 to 0.36) | <0.001 |  |  |
| <u>Model 3</u> |  |  | <0.001 | 5.44 | 0.7182 |
| MVPA | 0.05 | (0.04 to 0.06) | <0.001 |  |  |
| <u>Model 4</u> |  |  | <0.001 | 5.21 | 0.7417 |
| VPA | 0.30 | (0.24 to 0.36) | <0.001 |  |  |
| <u>Model 5</u> |  |  | <0.001 | 5.41 | 0.7215 |
| IG | 10.34 | (7.44 to 13.25) | <0.001 |  |  |
| TPA | 0.01 | (0.005 to 0.018) | <0.001 |  |  |
| <u>Model 6</u> |  |  | <0.001 | 5.19 | 0.7431 |
| VPA | 0.29 | (0.23 to 0.35) | <0.001 |  |  |
| TPA | 0.01 | (-0.00 to 0.01) | 0.059 |  |  |
| <u>Model 7</u> |  |  | <0.001 | 5.24 | 0.7394 |
| IG | 4.49 | (1.11 to 7.88) | 0.009 |  |  |
| AvAcc | 0.25 | (0.17 to 0.32) | <0.001 |  |  |
| *HEART* (n=67) |  |  |  |  |  |
| <u>Model 1</u> |  |  | <0.001 | 4.75 | 0.3802 |
| IG | 10.08 | (3.49 to 16.67) | 0.003 |  |  |
| <u>Model 2</u> |  |  | <0.001 | 4.87 | 0.3475 |
| AvAcc | 0.26 | (0.05 to 0.48) | 0.016 |  |  |
| <u>Model 3</u> |  |  | <0.001 | 4.73 | 0.3834 |
| MVPA | 0.05 | (0.02 to 0.09) | 0.003 |  |  |
| <u>Model 4</u> |  |  | <0.001 | 5.01 | 0.3082 |
| VPA | 0.43 | (-0.09 to 0.95) | 0.107 |  |  |
| <u>Model 5</u> |  |  | <0.001 | 4.76 | 0.3763 |
| IG | 9.33 | (2.04 to 16.96) | 0.009 |  |  |
| TPA | 0.01 | (-0.01 to 0.04) | 0.422 |  |  |
| <u>Model 6</u> |  |  | <0.001 | 5.00 | 0.3120 |
| VPA | 0.35 | (-0.19 to 0.89) | 0.203 |  |  |
| TPA | 0.01 | (-7.08 to 0.03) | 0.256 |  |  |
| <u>Model 7</u> |  |  | <0.001 | 4.73 | 0.3842 |
| IG | 7.85 | (2.41 to 15.46) | 0.043 |  |  |
| AvAcc | 0.14 | (-1.01 to 0.38) | 0.249 |  |  |

All models were adjusted for age \* sex + BMI (age and BMI were modelled using restricted cubic splines). Abbreviations: AvAcc, average acceleration; HEALTHY, healthy individuals; HEART, patients with heart failure; IG, intensity gradient; MVPA, moderate to vigorous physical activity; TPA, time in physical activity; VPA, vigorous physical activity.

When combining IG as a metric of PA intensity and TPA as a surrogate of PA time (Model 5), both metrics were independent predictors of VO_2peak_. However, when including VPA and TPA (model 6), there was little evidence for the latter being an independent predictor of VO_2peak_. In HEART, there was little evidence for a difference in fit between model 1 (IG), 2 (AvAcc), or 7 (IG+AvAcc). Both IG and AvAcc were associated with VO_2peak_ with only IG being independent of AvAcc but not vice versa. Moreover, the best model with cut-point-based metric (model 3 with MVPA) performed equal to models 1 (IG) and 7 (AvAcc+IG), but was superior to model 2 (AvAcc) (p=0.012). Adding TPA to the model with IG or VPA did not significantly improve model fits (models 5&6).

### Reference values for healthy adults

Sex-specific percentile curves were created for IG, AvAcc (see Figure 2), MVPA, and VPA (see Supplementary Figure 3). Empirical data for IG and AvAcc per age decade are presented in Table 3. Data for MVPA and VPA are shown in Supplementary Table 2.

**Figure 2.**
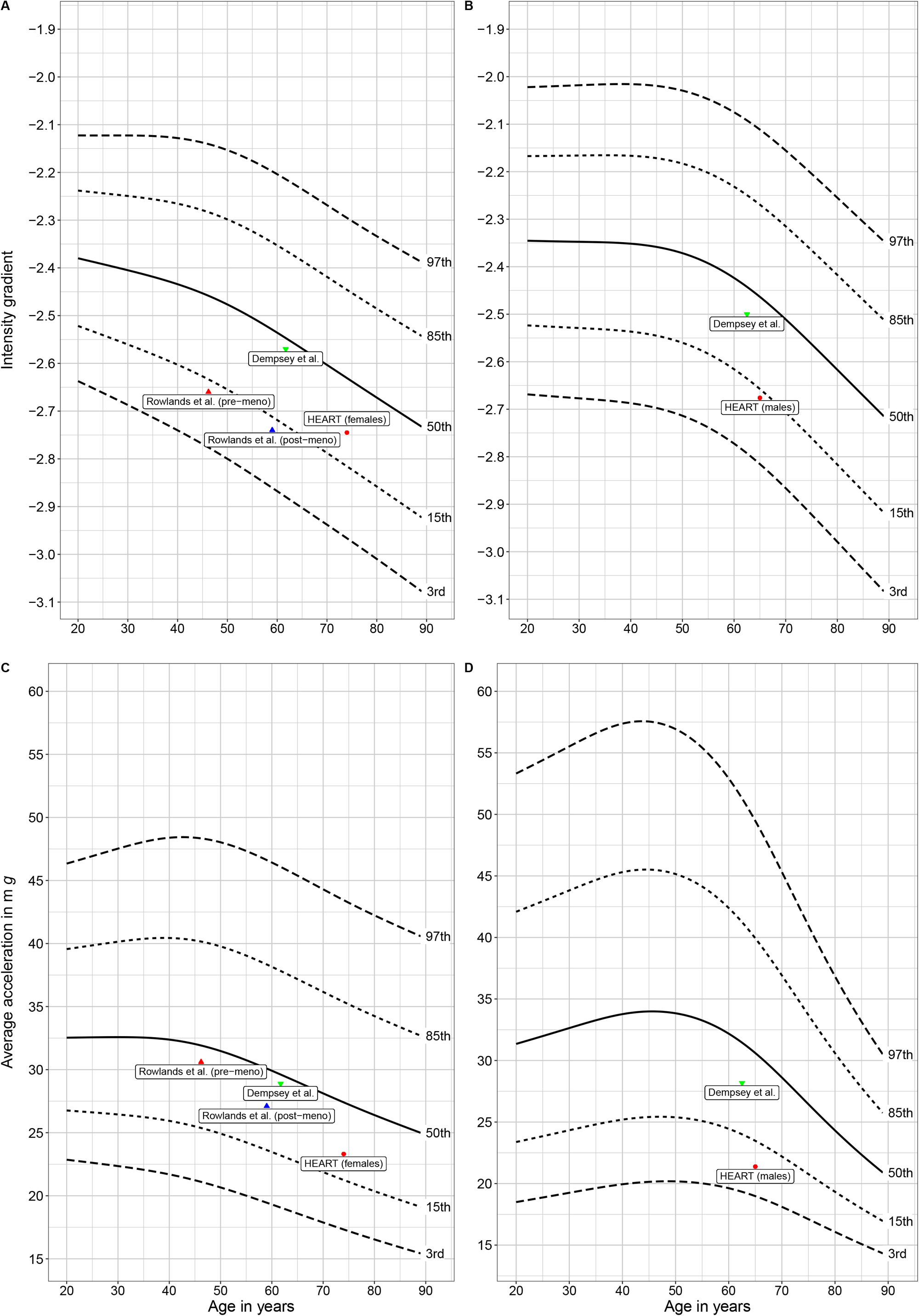
Percentile curves for intensity gradient (A & B) and average acceleration (C & D) in relation to age in healthy adults. The left column shows data for females and the right column for males. Mean values of IG and AvAcc of other cohorts were plotted to highlight the potential of the reference curves for comparability across studies and between devices. Data by Rowlands et al. (1): pre- and post-menopausal women (n_pre_=1218, n_post_=1316; device: Axivity). Dempsey et al. (23): participants of the UK Biobank study (females, n = 51,509; males, n=36,903, device: Axivity, dominant wrist). Our data of patients with heart failure (HEART; device: GENEActiv) were also plotted.

**Table 3.**
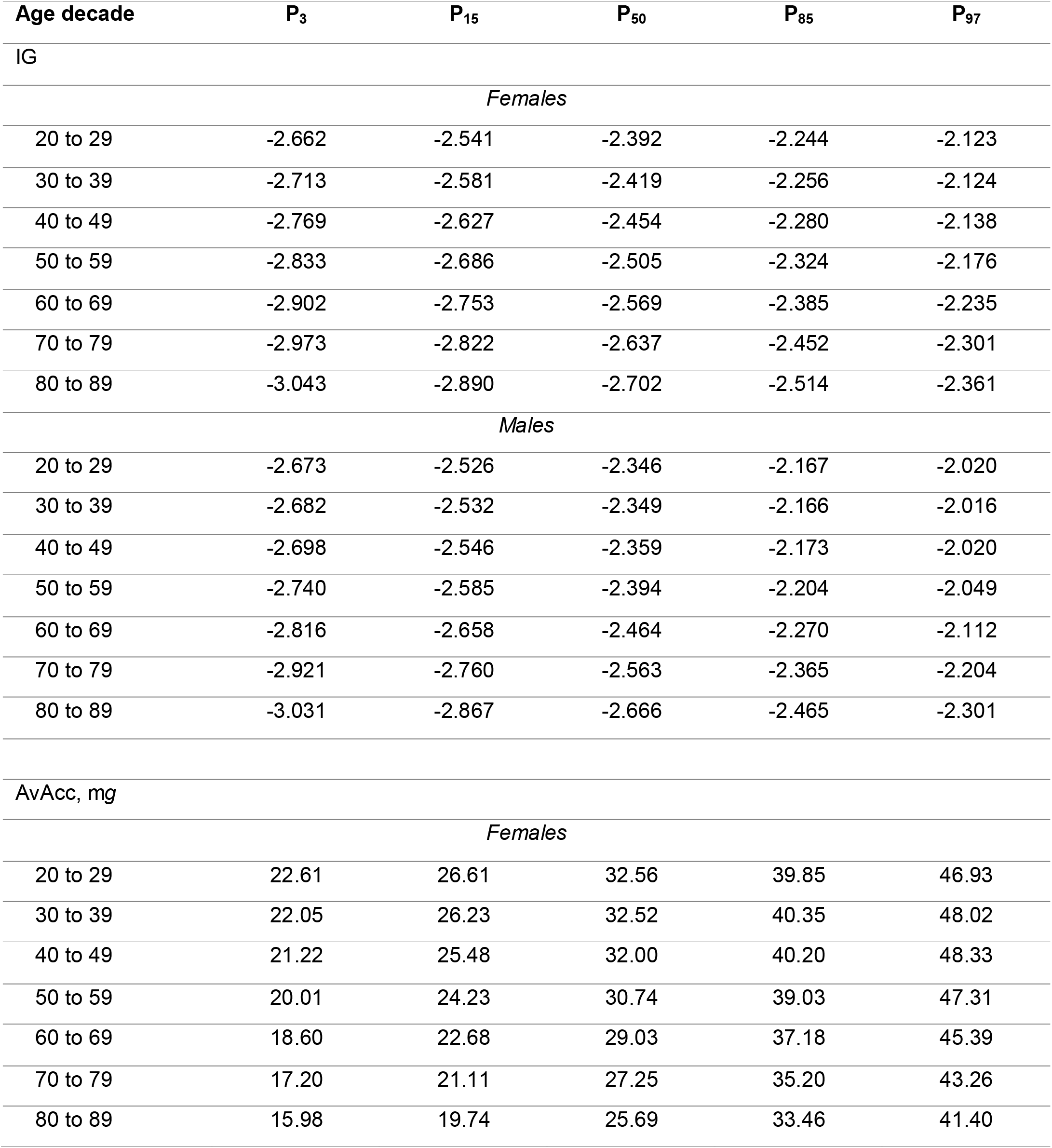
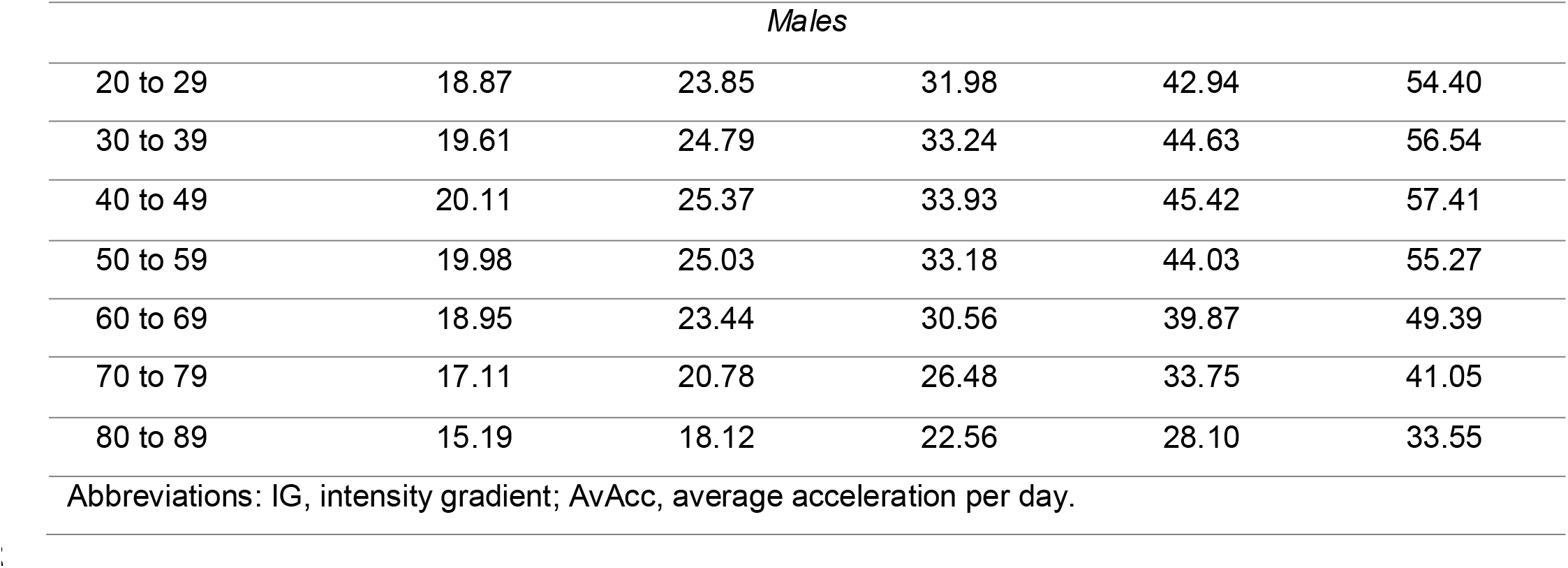
Empirical data for IG and AvAcc in healthy individuals stratified by age decade and sex.

| Age decade | P<sub>3</sub> | P<sub>15</sub> | P<sub>50</sub> | P<sub>85</sub> | P<sub>97</sub> |
| --- | --- | --- | --- | --- | --- |
| IG |  |  |  |  |  |
| *Females* |  |  |  |  |  |
| 20 to 29 | -2.662 | -2.541 | -2.392 | -2.244 | -2.123 |
| 30 to 39 | -2.713 | -2.581 | -2.419 | -2.256 | -2.124 |
| 40 to 49 | -2.769 | -2.627 | -2.454 | -2.280 | -2.138 |
| 50 to 59 | -2.833 | -2.686 | -2.505 | -2.324 | -2.176 |
| 60 to 69 | -2.902 | -2.753 | -2.569 | -2.385 | -2.235 |
| 70 to 79 | -2.973 | -2.822 | -2.637 | -2.452 | -2.301 |
| 80 to 89 | -3.043 | -2.890 | -2.702 | -2.514 | -2.361 |
| *Males* |  |  |  |  |  |
| 20 to 29 | -2.673 | -2.526 | -2.346 | -2.167 | -2.020 |
| 30 to 39 | -2.682 | -2.532 | -2.349 | -2.166 | -2.016 |
| 40 to 49 | -2.698 | -2.546 | -2.359 | -2.173 | -2.020 |
| 50 to 59 | -2.740 | -2.585 | -2.394 | -2.204 | -2.049 |
| 60 to 69 | -2.816 | -2.658 | -2.464 | -2.270 | -2.112 |
| 70 to 79 | -2.921 | -2.760 | -2.563 | -2.365 | -2.204 |
| 80 to 89 | -3.031 | -2.867 | -2.666 | -2.465 | -2.301 |
| AvAcc, mg |  |  |  |  |  |
| *Females* |  |  |  |  |  |
| 20 to 29 | 22.61 | 26.61 | 32.56 | 39.85 | 46.93 |
| 30 to 39 | 22.05 | 26.23 | 32.52 | 40.35 | 48.02 |
| 40 to 49 | 21.22 | 25.48 | 32.00 | 40.20 | 48.33 |
| 50 to 59 | 20.01 | 24.23 | 30.74 | 39.03 | 47.31 |
| 60 to 69 | 18.60 | 22.68 | 29.03 | 37.18 | 45.39 |
| 70 to 79 | 17.20 | 21.11 | 27.25 | 35.20 | 43.26 |
| 80 to 89 | 15.98 | 19.74 | 25.69 | 33.46 | 41.40 |

| *Males* |  |  |  |  |  |
| --- | --- | --- | --- | --- | --- |
| 20 to 29 | 18.87 | 23.85 | 31.98 | 42.94 | 54.40 |
| 30 to 39 | 19.61 | 24.79 | 33.24 | 44.63 | 56.54 |
| 40 to 49 | 20.11 | 25.37 | 33.93 | 45.42 | 57.41 |
| 50 to 59 | 19.98 | 25.03 | 33.18 | 44.03 | 55.27 |
| 60 to 69 | 18.95 | 23.44 | 30.56 | 39.87 | 49.39 |
| 70 to 79 | 17.11 | 20.78 | 26.48 | 33.75 | 41.05 |
| 80 to 89 | 15.19 | 18.12 | 22.56 | 28.10 | 33.55 |
| Abbreviations: IG, intensity gradient; AvAcc, average acceleration per day. |  |  |  |  |  |

\*\*\* insert Figure 2 here \*\*\*

CRF-specific percentile curves for IG and AvAcc are available in Figure 3.

**Figure 3.**
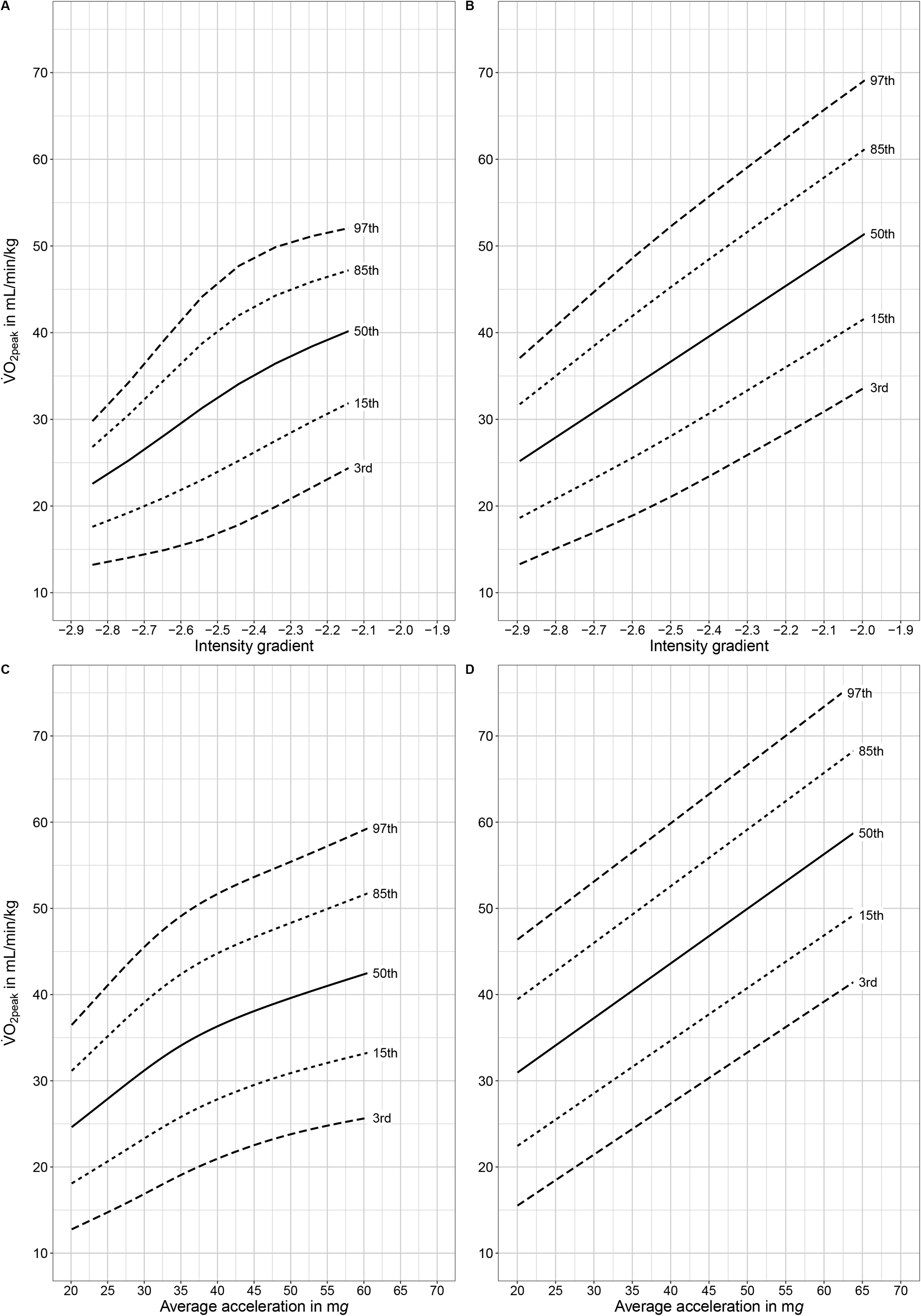
Percentile curves for intensity gradient (A & B) and average acceleration (C & D) in relation to VO_2peak_ in healthy adults stratified by sex. The left column shows data for females and the right column for males. Abbreviations: VO_2peak_, peak oxygen uptake.

We developed the application ‘rawacceleration’ using these reference data. It is freely available on GitHub (https://github.com/FSchwendinger/rawacceleration). Further insights into ‘rawacceleration’ are given in the Supplement and discussion section.

## DISCUSSION

The major novel findings are IG and AvAcc yield complementary information on PA, particularly intensity, with both being important predictors of VO_2peak_ in healthy individuals and patients with heart failure. Both metrics together had similar predictive value for VO_2peak_ as the best cut-point-based metric in healthy adults. In patients with heart failure, both IG alone and IG+AvAcc together had similar predictive value as the best cut-point-based metric with IG being independent of AvAcc. Cut-point-free metrics also have the advantage of being comparable across cohorts and the most commonly used accelerometers.[6, 24] Importantly, this population-based study produced percentile curves and reference values for both cut-point-free and traditional accelerometer metrics for healthy adults aged 20 to 89 years. This addresses the limited interpretability of the AvAcc and IG metrics by placing them into context. Our results provide generalised support for policy making.

### Inter-relationship between accelerometer metrics and VO_2peak_ in HEALTHY and HEART

We confirmed previous findings in children and adults with type 2 diabetes suggesting AvAcc and IG provide a complementary picture of PA.[6, 7] Our study demonstrated this in healthy adults and patients with heart failure by examining correlations between accelerometer metrics. These findings are strengthened by our explorative principle component analysis. Lower communalities were seen for IG than for AvAcc or MVPA indicating IG may yield a perspective on PA not covered by other metrics. AvAcc and IG may thus provide a more complete impression of an individual’s PA profile without having to rely on accelerometer cut-points.

AvAcc and IG have been described to reflect PA volume and intensity, respectively.[6] Yet, correlations between TPA, VPA, IG, and AvAcc indicate that AvAcc conveys information on both PA intensity and volume. It should therefore be noted that cut-point-free metrics allow the further investigation of intensity/volume, despite their overlap, as IG captures the distribution as well as quantity of intensity. This is more difficult with cut-point-based metrics because of their categorical and population-dependent nature. Thus AvAcc and IG may be particularly appropriate when attempting to differentiate between the contribution of PA intensity and volume to CRF as well as other health parameters (e.g. body fatness, BMI, physical function).[6, 7]

IG and AvAcc together had the greatest predictive value for VO_2peak_ in healthy adults and both were independently associated, explaining a unique fraction of the variance. This is partly compliant with a study in children that estimated CRF from a shuttle-run-test.[7] IG or AvAcc as the only independent variable in the adjusted regression models was significantly associated with estimated CRF.[7] Yet, only IG remained significantly associated with CRF when AvAcc was added to the model.[7] AvAcc may consequently be more relevant in adults than in children. This is possibly due to different activity patterns with higher IG and AvAcc together with less sedentary time in children.[7] This highlights the utility of combining these two metrics to gain insight into the relative importance of volume and intensity of the PA profile which differs across populations.[6]

Interestingly, TPA significantly added to model 5 also containing IG. Both intensity distribution (IG) and time spent in PA (TPA) may thus be relevant for VO_2peak_. Notably, AvAcc captures both time and intensity of PA, thus reflecting activity volume, while IG captures the distribution of the intensity of activity. Combining IG and AvAcc thus yields complementary perspectives on PA intensity.

In patients with heart failure, cut-point-free metrics had similar predictive value for VO_2peak_ as the best cut-point-based metric, MVPA. Yet, unlike in healthy adults, only IG was independent of AvAcc but not vice versa. This may reflect relatively little activity of higher intensity in these patients; it is in contrast with findings of Dawkins et al. (25) and suggests that a focus on intensity of activity, i.e. increasing the breadth of the intensity distribution, may be more important than volume if intervening in this population.

This study demonstrated that IG and AvAcc are not only potentially more robust accelerometer metrics than MVPA, VPA, or TPA, as they do not rely on cut-points, but also have similar predictive value for VO_2peak_ and, in turn, indirectly for the risk of mortality and longevity.[8, 26] Indeed, Dempsey et al. (23) recently demonstrated that AvAcc and IG were independently associated with incident cardiovascular disease in participants from UK Biobank. These results and those of previous studies[6, 7] provide an evidence-based rationale that cut-point-free accelerometer metrics facilitate capture of the volume and intensity distribution of the PA profile across populations, and thus may be a viable alternative to cut-point-based outcomes in the measurement of PA.

### Reference values for healthy adults

Our reference values and percentile curves may be pivotal to assessing the appropriateness of PA on population-or individual-level and to determining the level of PA necessary for a healthy lifestyle.[6]

IG and AvAcc at the 50^th^ percentile are higher than in participants of the UK Biobank Study aged 40 to 69 years, despite them wearing the accelerometer on their dominant hand which typically elicits accelerations approximately 10% higher.[1, 27] This seems reasonable in light of the good health condition (see inclusion/exclusion criteria) and the likely higher CRF[12] of the COmPLETE cohort compared to the aforementioned study. Hence, our reference values might be desirable benchmarks for healthy PA patterns in adults.

Age trajectories of IG and AvAcc were curvilinear and inverse for both sexes (see Figure 2). This is in line with the accelerometer data of another Swiss adult cohort.[28] Interestingly, the decline in IG and AvAcc with age seems to resemble that of CRF and muscular strength in the COmPLETE cohort.[12, 29] With age, less time is spent at higher absolute intensities which might be due to reduced physical functioning as apparent by lower CRF and muscular strength. However, when expressed in relative terms (as % of an individual’s VO_2peak_), intensity may still be high. This explanation is underpinned by questionnaire data, which may also reflect the rate of perceived exertion, showing more moderate and vigorous PA in individuals aged > 59 years than in 40 to 59-year-olds.[28] Moreover, the inverse relationship seen between accelerometer-based VPA and age seems not to be apparent in questionnaire data.[28]

The proposed reference values may also be useful when PA is measured with other raw-acceleration, wrist-worn accelerometers. Migueles et al. (24) found high inter-instrument reliability for four common accelerometers (GENEActiv, Movisens Move 4, ActiGraph GT3X+, and Axivity AX3) provided that raw data are processed identically. The between-device difference ranged between 1 (95% CI: -6 to 7) and 8 (95% CI: 1 to 15) min per day.[24] Our reference values may thus be applicable for at least the examined accelerometers (i.e. GENEActiv, Movisens Move 4, ActiGraph GT3X+, and Axivity AX3).[24]

Reference values and percentile curves for cut-point-based metrics (VPA and MVPA, see Supplement: Figure 3 and Table 2) were also created. These are still commonly used and directly interpretable. Comparison to these data is however only possible when using the cut-points by Hildebrand et al. (3) (MVPA=0.1 *g* and VPA=0.4 *g*) and processing data identically.[24] Due to the aforementioned disadvantages of cut-point-based accelerometer metrics (also discussed elsewhere[5]), cut-point-free metrics should be reported in future studies examining PA with raw-acceleration accelerometers.

Importantly, the reference values for traditional metrics are specific to accelerometer data and examine the whole activity profile. They are not comparable to e.g. the PA guidelines by the WHO which focus on MVPA and were developed predominantly from self-reported data.[2, 30] This is supported by numerous studies.[31, 32]

Our reference values facilitate interpretation of accelerometer-assessed PA data relative to a ‘healthy norm’, and may thus be an important complement to the current PA guidelines by the WHO.[30]

### Limitations

PA levels differ between countries.[33] Nonetheless, it seems to be desirable to promote the present reference values also in countries other than Switzerland considering the good general health of our cohort in combination with their high CRF.[12] Importantly, the present reference values are only applicable if the data are processed similarly (see Methods section and configuration file). The R-package GGIR will facilitate this process.[10] Large differences in sampling frequency of accelerometer data may impact outcomes. In a study comparing 25 Hz with 100 Hz, the lower sampling frequency led to 3.1%–13.9% lower overall acceleration values.[34] The difference may however be smaller at 50 Hz, as used in our study. Yet, sampling frequency should be considered when comparing PA metrics to the present reference values. Available algorithms may help transform PA metrics to match those obtained from higher sampling frequencies.[34] Lastly, no causation between PA and VO_2peak_ can be implied from the present study.

## Data Availability

All data produced in the present study are available upon reasonable request to the authors

## CLINICAL IMPLICATIONS

IG and AvAcc provide complementary information on PA, particularly the intensity distribution of the PA profile. IG and AvAcc were strongly associated with VO_2peak_ in healthy adults and patients with heart failure. These metrics are independent of accelerometer cut-points, facilitate investigation of the relative contribution of intensity and volume of PA for a given health marker, and thus may be important parameters with regard to general health and risk for mortality and non-communicable diseases. To the best of our knowledge, this is the first study to produce reference values for IG and AvAcc for healthy adults. These reference values may be valuable to judge an individual’s or a population’s level of PA and facilitate cross-study comparison. The ‘rawacceleration’ application may be helpful for to compare individual-or cohort-level data to healthy age- and sex-matched adults and translate results into meaningful outcomes in research settings and clinical practice.

## Declarations

### Ethics approval and consent to participate

This study was approved by the Ethics Committee of North-western and Central Switzerland (EKNZ 2017–01451). All participants in this study provided informed written consent.

### Consent for publication

Not applicable.

### Availability of data and materials

The datasets used and/or analysed during the current study are available from the corresponding author on reasonable request.

### Authors contributions

FS conceptualised the manuscript and wrote the original draft. FS and DI analysed and interpreted the data. JW, AST, TH, and RK conceptualised the study and defined the methods. JW and RK collected the data. FS, JW, and RK were responsible for data curation. JW, DI, RK, AVR, TH, and AST revised the manuscript. AST obtained funding for the project. All authors approved the final version of the manuscript.

## Acknowledgements

We are grateful for the valuable contributions of all study participants in the COmPLETE study.

## List of abbreviations

AvAcc: Average acceleration
CRF: Cardiorespiratory fitness
ENMO: Euclidean norm minus one
HEALTHY: Healthy adults
HEART: Patients with heart failure
IG: Intensity gradient
MVPA: Moderate-to-vigorous physical activity
NT-proBNP: N-terminal pro b-type natriuretic peptide
NYHA: New York Heart Association
PA: Physical activity
SD: Standard deviation
SED: Sedentary time
TPA: Total physical activity
VO_2peak_: Peak oxygen uptake
VPA: Vigorous physical activity

## REFERENCES

1. Rowlands AV, Fairclough SJ, Yates TOM, Edwardson CL, Davies M, Munir F, et al. Activity Intensity, Volume, and Norms: Utility and Interpretation of Accelerometer Metrics. Med Sci Sports Exerc. 2019;51(11):2410–22.

2. Troiano RP, McClain JJ, Brychta RJ, Chen KY. Evolution of accelerometer methods for physical activity research. Br J Sports Med. 2014;48(13):1019–23.

3. Hildebrand M, van Hees VT, Hansen BH, Ekelund U. Age Group Comparability of Raw Accelerometer Output from Wrist- and Hip-Worn Monitors. Med Sci Sports Exerc. 2014;46(9):1816–24.

4. Esliger DW, Rowlands AV, Hurst TL, Catt M, Murray P, Eston RG. Validation of the GENEA Accelerometer. Med Sci Sports Exerc. 2011;43(6):1085–93.

5. Schwendinger F, Wagner J, Infanger D, Schmidt-Trucksäss A, Knaier R. Methodological aspects for accelerometer-based assessment of physical activity in heart failure and health. BMC Med Res Methodol. 2021;21(1):251.

6. Rowlands AV, Edwardson CL, Davies MJ, Khunti K, Harrington DM, Yates T. Beyond Cut Points: Accelerometer Metrics that Capture the Physical Activity Profile. Med Sci Sports Exerc. 2018;50(6):1323–32.

7. Fairclough SJ, Taylor S, Rowlands AV, Boddy LM, Noonan RJ. Average acceleration and intensity gradient of primary school children and associations with indicators of health and well-being. J Sports Sci. 2019;37(18):2159–67.

8. Kodama S, Saito K, Tanaka S, Maki M, Yachi Y, Asumi M, et al. Cardiorespiratory Fitness as a Quantitative Predictor of All-Cause Mortality and Cardiovascular Events in Healthy Men and Women: A Meta-analysis. JAMA. 2009;301(19):2024–35.

9. Wagner J, Knaier R, Infanger D, Arbeev K, Briel M, Dieterle T, et al. Functional aging in health and heart failure: the COmPLETE Study. BMC Cardiovasc Disord. 2019;19(1):180.

10. Migueles JH, Rowlands AV, Huber F, Sabia S, van Hees VT. GGIR: A Research Community–Driven Open Source R Package for Generating Physical Activity and Sleep Outcomes From Multi-Day Raw Accelerometer Data. J Meas Phys Behav. 2019;2(3):188–96.

11. van Hees VT, Gorzelniak L, Dean León EC, Eder M, Pias M, Taherian S, et al. Separating Movement and Gravity Components in an Acceleration Signal and Implications for the Assessment of Human Daily Physical Activity. PloS ONE. 2013;8(4):e61691. 10.1371/journal.pone.0061691.

12. Wagner J, Knaier R, Infanger D, Königstein K, Klenk C, Carrard J, et al. Novel CPET Reference Values in Healthy Adults: Associations with Physical Activity. Med Sci Sports Exerc. 2020;53(1):26–37.

13. R Core Team. R: A language and environment for statistical computing Vienna, Austria: R Foundation for Statistical Computing; 2021 [Available from: https://www.R-project.org/].

14. Kassambara A, Mundt F. Factoextra: Extract and Visualize the Results of Multivariate Data Analyses. R Package Version 1.0.7. 2020.

15. Mevik B-H, Wehrens R. The pls Package: Principal Component and Partial Least Squares Regression in R. J Stat Softw. 2007;18(2):1–23.

16. Harrell FE, Jr. Regression modeling strategies. 2 ed: Springer International Publishing; 2015.

17. Vuong QH. Likelihood Ratio Tests for Model Selection and Non-Nested Hypotheses. Econometrica. 1989;57(2):307–33.

18. Stasinopoulos DM, Rigby RA. Generalized Additive Models for Location Scale and Shape (GAMLSS) in R. J Stat Softw. 2007;23(7):1–46.

19. Stasinopoulos MD, Rigby, R.A., Heller, G.Z., Voudouris, V., De Bastiani, F.. Flexible Regression and Smoothing: Using GAMLSS in R. 1 ed: Chapman and Hall/CRC; 2017.

20. Eilers PHC, Marx, B. D. Practical smoothing. The joys of P-splines: Cambridge University Press; 2021.

21. van Buuren S, Fredriks M. Worm plot: a simple diagnostic device for modelling growth reference curves. Stat Med. 2001;20(8):1259–77.

22. Chang W, Cheng J, Allaire J, Xie Y, McPherson J. Shiny: web application framework for R. R package version. 2017;1(5):2017.

23. Dempsey PC, Rowlands AV, Strain T, Zaccardi F, Dawkins NP, Razieh C, et al. Physical activity volume, intensity and incident cardiovascular disease. Eur Heart J. 2022:43(46):4789–4800.

24. Migueles JH, Molina-Garcia P, Torres-Lopez LV, Cadenas-Sanchez C, Rowlands AV, Ebner-Priemer UW, et al. Equivalency of four research-grade movement sensors to assess movement behaviors and its implications for population surveillance. Sci Rep. 2022;12(1):5525. 10.1038/s41598-022-09469-2.

25. Dawkins NP, Yates T, Edwardson CL, Maylor B, Henson J, Hall AP, et al. Importance of Overall Activity and Intensity of Activity for Cardiometabolic Risk in Those with and Without a Chronic Disease. Med Sci Sports Exerc. 2022;54(9):1582–90.

26. Mok A, Khaw K-T, Luben R, Wareham N, Brage S. Physical activity trajectories and mortality: population based cohort study. BMJ. 2019;365:2323. 10.1136/bmj.l2323.

27. Buchan DS, Boddy LM, McLellan G. Comparison of Free-Living and Laboratory Activity Outcomes from ActiGraph Accelerometers Worn on the Dominant and Non-Dominant Wrists. Meas Phys Edu Exerc Sci. 2020;24(4):247–57.

28. Wanner M, Hartmann C, Pestoni G, Martin BW, Siegrist M, Martin-Diener E. Validation of the Global Physical Activity Questionnaire for self-administration in a European context. BMJ Open Sport Exerc Med. 2017;3(1):e000206. 10.1136/bmjsem-2016-000206.

29. Lichtenstein E, Wagner J, Knaier R, Infanger D, Roth R, Hinrichs T, et al. Norm Values of Muscular Strength Across the Life Span in a Healthy Swiss Population: The COmPLETE Study. Sports Health. 2022. 10.1177/19417381221116345.

30. Bull FC, Al-Ansari SS, Biddle S, Borodulin K, Buman MP, Cardon G, et al. World Health Organization 2020 guidelines on physical activity and sedentary behaviour. Br J Sports Med. 2020;54(24):1451–62.

31. Tucker JM, Welk GJ, Beyler NK. Physical activity in U.S.: adults compliance with the Physical Activity Guidelines for Americans. Am J Prev Med. 2011;40(4):454–61.

32. Schuna JM, Jr., Johnson WD, Tudor-Locke C. Adult self-reported and objectively monitored physical activity and sedentary behavior: NHANES 2005-2006. Int J Behav Nutr Phys Act. 2013;10:126. 10.1186/1479-5868-10-126.

33. Bauman A, Ainsworth BE, Sallis JF, Hagströmer M, Craig CL, Bull FC, et al. The Descriptive Epidemiology of Sitting: A 20-Country Comparison Using the International Physical Activity Questionnaire (IPAQ). Am J Prev Med. 2011;41(2):228–35.

34. Small S, Khalid S, Dhiman P, Chan S, Jackson D, Doherty A, et al. Impact of Reduced Sampling Rate on Accelerometer-Based Physical Activity Monitoring and Machine Learning Activity Classification. J Meas Phys Behav. 2021;4(4):298–310.

