## Supplement for "Reference values for cut-point-free and traditional accelerometer metrics and associations with cardiorespiratory fitness: a cross-sectional study of healthy adults aged 20 to 89 years"

**Supplementary material**

**Accelerometer data processing**

Raw data processing was done using the GGIR package version 2.6-0[1, 2] in R (R Foundation for Statistical Computing, Vienna, Austria).[3] This included auto-calibration using local gravity as a reference[4] and sleep detection differentiating between waking time and presumed sleeping time.[5] Calibration errors > 0.01 g led to exclusion.[4] Moreover, GGIR computed Euclidean norm minus one (ENMO) averaged over 5 s in m*g*.[6] The GGIR configuration file is available as Supplement 1. All PA data refer to the time from midnight to midnight (24 h). Acceleration cut-points of 0.03, 0.1, and 0.4 *g* were applied to categorise PA intensity into light (≥ 2 METs), moderate (≥ 3 METs), and vigorous (≥ 6 METs), respectively.[6, 7] Moderate-to-vigorous PA (MVPA) is the sum of moderate and vigorous PA (VPA). IG is computed as the natural log relationship of intensity, quantified by the midrange of incremental acceleration bins with size 25 m*g* (e.g. 25 m*g* bin=12.5), and time accumulated in each acceleration bin.[8, 9] It reflects the drop of time accumulated in increasing intensity bins and is always negative.[8, 9] PA data were averaged across all days without weighting. All GGIR variables used in the analyses are reported below.

**GGIR variable names**

|  | Name in GGIR | Name in manuscript |
| --- | --- | --- |
| Part 2 | AD_ig_gradient_ENMO_0.24hr | Intensity gradient |
|  | AD_mean_ENMO_mg_0.24hr | Average acceleration |
|  | AD_ig_intercept_ENMO_0.24hr | Intercept of intensity gradient regression line |
|  | AD_ig_rsquared_ENMO_0.24hr | R^2^ of intensity gradient regression line |
| Part 5 | dur_day_total_IN_min_pla | Sedentary time |
|  | dur_day_total_LIG_min_pla | Light physical activity |
|  | dur_day_total_MOD_min_pla | Moderate physical activity |
|  | dur_day_total_VIG_min_pla | Vigorous physical activity |

**Explorative principal component analysis**

In healthy individuals (HEALTHY), the first component of the analysis accounted for 59.7% of the variation of all included variables. The highest loadings for component 1 were seen for average acceleration (AvAcc), moderate-to-vigorous physical activity (MVPA), vigorous PA (VPA), intensity gradient (IG), time in physical activity (TPA) (0.48, 0.45, 0.38, 0.35, respectively). This indicates that these variables reflect some similar information and thus have mild to moderate redundancy. The calculation of communalities showed that component 1 explained 94.7%, 83.9%, 61.5%, 51.2%, and 48.0% of the variance in AvAcc, MVPA, VPA, IG, and TPA, respectively.

In patients with heart failure (HEART), component 1 accounted for 57.3% of the variation of all included variables. The highest loadings occurred for AvAcc, MVPA, TPA, IG, and VPA (0.48, 0.47, 0.42, 0.34, 0.31, respectively). Component 1 explained 93.4%, 87.6%, 71.0%, 45.5%, and 39.2% of the variance in AvAcc, MVPA, TPA, IG, and VPA, respectively.

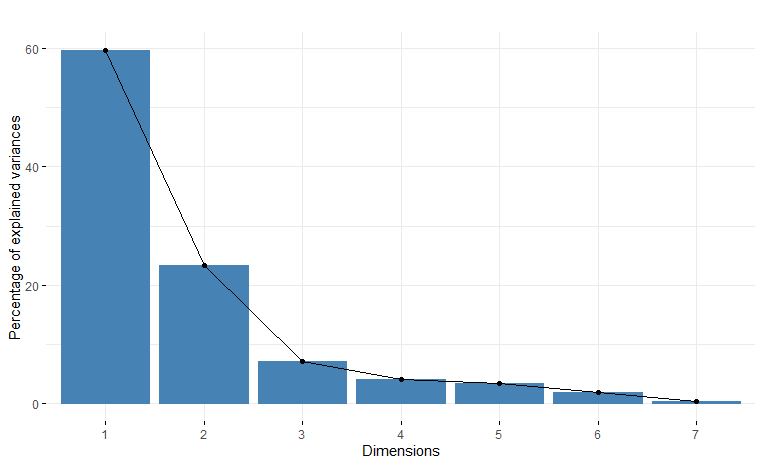
**Supplementary Figure 1.** Scree plot displaying the results of the principal component analysis in healthy individuals.

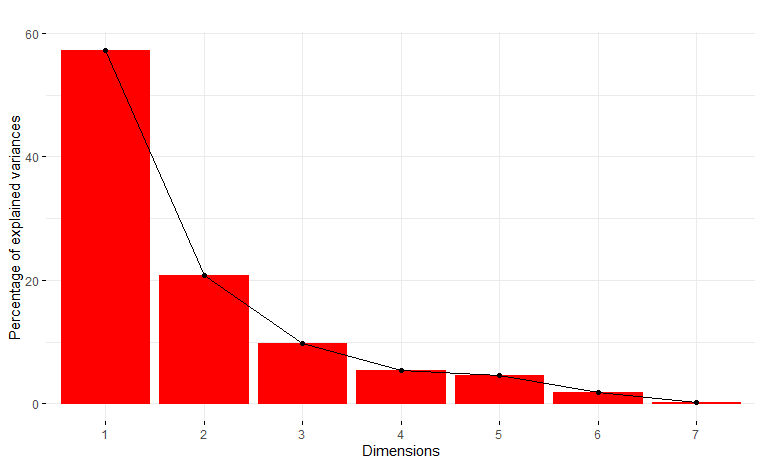
**Supplementary Figure 2.** Scree plot displaying the results of the principal component analysis in patients with heart failure.

**Partial least squares analyses**

Supplementary table 1 shows the results of the partial least squares analyses for healthy individuals and patients with heart failure, respectively.

| Supplementary Table 1. Partial least squares analyses for HEALTHY and HEART with peak oxygen uptake as the dependent variable. | | | | |
| --- | --- | --- | --- | --- |
|  | **HEALTHY** | | **HEART** | |
| Parameters | **Coefficient** | **Variance explained** | **Coefficient** | **Variance explained** |
| V̇O_2peak_ | - | 35.92% | - | 22.23% |
| IG | 1.85 | - | 0.89 | - |
| AvAcc | 1.57 | - | 0.66 | - |
| VPA | 1.61 | - | 0.65 | - |
| MVPA | 1.54 | - | 0.75 | - |
| TPA | 0.38 | - | 0.34 | - |
| SED | -0.33 | - | -0.24 | - |
| Abbreviations: AvAcc, average acceleration; HEALTHY, healthy individuals; HEART, patients with heart failure; IG, intensity gradient; MVPA, moderate-to-vigorous physical activity; SED, sedentary time; TPA, time in physical activity; V̇O_2peak_, peak oxygen uptake; VPA, vigorous physical activity. | | | | |

IG and AvAcc followed by VPA and MVPA were the most important predictors for V̇O_2peak_ in HEALTHY. IG, MVPA, AvAcc, and VPA seemed to be the most important predictors in HEART.

**Reference values for moderate- to vigorous and vigorous physical activity**

Sex-specific percentile curves for MVPA and VPA are available in Supplementary Figure 3. Empirical data for these parameters per age decade are presented in Supplementary Table 2.

| Supplementary Table 2. Empirical data for MVPA and VPA in healthy individuals stratified by age decade and sex. | | | | | |
| --- | --- | --- | --- | --- | --- |
| Age decade | **P_3_** | **P_15_** | **P_50_** | **P_85_** | **P_97_** |
| MVPA, min^.^day^-1^ | | | | | |
| *Females* | | | | | |
| 20 to 29 | 82 | 102 | 132 | 173 | 214 |
| 30 to 39 | 75 | 95 | 127 | 170 | 216 |
| 40 to 49 | 67 | 87 | 120 | 166 | 216 |
| 50 to 59 | 58 | 78 | 111 | 158 | 211 |
| 60 to 69 | 49 | 68 | 100 | 148 | 204 |
| 70 to 79 | 41 | 58 | 90 | 138 | 196 |
| 80 to 89 | 34 | 50 | 80 | 129 | 190 |
| *Males* | | | | | |
| 20 to 29 | 64 | 88 | 125 | 171 | 216 |
| 30 to 39 | 65 | 91 | 132 | 183 | 234 |
| 40 to 49 | 65 | 92 | 135 | 190 | 244 |
| 50 to 59 | 60 | 87 | 130 | 185 | 240 |
| 60 to 69 | 50 | 74 | 113 | 163 | 213 |
| 70 to 79 | 38 | 57 | 88 | 130 | 171 |
| 80 to 89 | 28 | 42 | 67 | 99 | 132 |
| VPA, min^.^day^-1^ | | | | | |
| *Females* | | | | | |
| 20 to 29 | 0.8 | 1.9 | 5.5 | 15.6 | 36.3 |
| 30 to 39 | 0.8 | 1.9 | 5.5 | 15.9 | 38.0 |
| 40 to 49 | 0.7 | 1.6 | 4.9 | 14.8 | 36.2 |
| 50 to 59 | 0.5 | 1.2 | 3.6 | 11.2 | 28.1 |
| 60 to 69 | 0.3 | 0.7 | 2.3 | 7.3 | 18.9 |
| 70 to 79 | 0.2 | 0.4 | 1.4 | 4.7 | 12.3 |
| 80 to 89 | 0.1 | 0.3 | 0.9 | 3.0 | 8.2 |
| *Males* | | | | | |
| 20 to 29 | 0.3 | 1.5 | 6.3 | 17.3 | 31.9 |
| 30 to 39 | 0.3 | 1.7 | 7.3 | 20.0 | 37.0 |
| 40 to 49 | 0.4 | 2.0 | 8.5 | 23.3 | 43.1 |
| 50 to 59 | 0.4 | 2.0 | 8.5 | 23.1 | 42.8 |
| 60 to 69 | 0.3 | 1.4 | 6.0 | 16.5 | 30.5 |
| 70 to 79 | 0.1 | 0.7 | 2.9 | 7.8 | 14.5 |
| 80 to 89 | 0.0 | 0.3 | 1.1 | 3.1 | 5.7 |
| Abbreviations: MVPA, moderate-to-vigorous physical activity; VPA, vigorous physical activity. | | | | | |

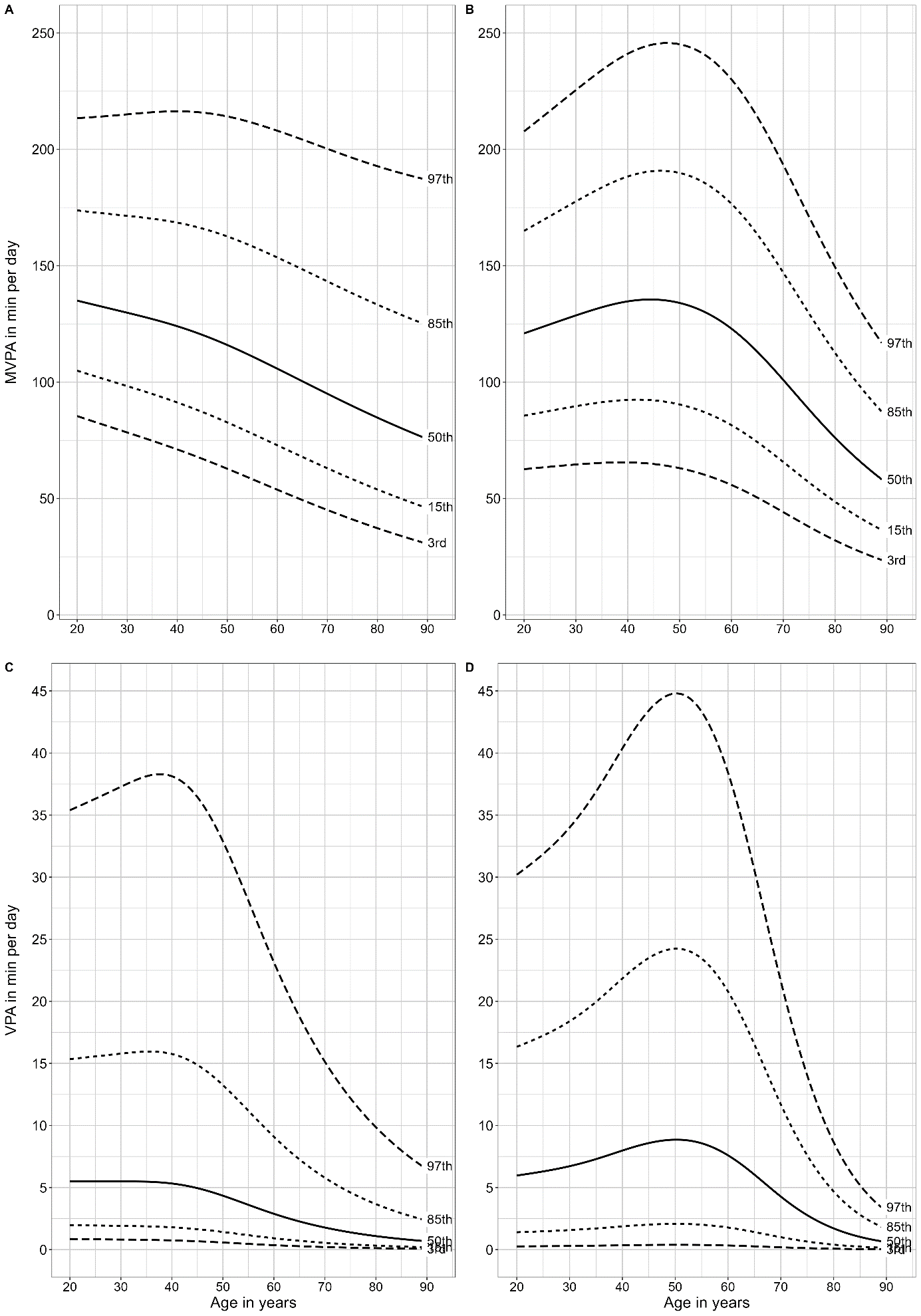

**Supplementary Figure 3.** Percentile curves for moderate-to-vigorous physical activity (A & B) and vigorous physical activity (C & D) in relation to age in healthy Swiss adults. The left column shows data for females and the right column for males. Abbreviations: MVPA, moderate-to-vigorous physical activity; VPA, vigorous physical activity.

**Preview of the new application ‘rawacceleration’**

The new application provides the user with several data entry options. Data of individuals (see Supplementary Figure 4), aggregated cohort-level data stratified by sex or non-stratified (cohort means/medians), or raw-level data can be entered. Once all required data have been entered and the calculations have been triggered, the results will be provided in a separate panel (panel 2: View results). For the first two data entry options, our percentile curves presented in the manuscript will be plotted. The user’s data will be shown in the plot (shown in green in Supplementary Figure 5) together with the exact percentiles. In another panel (panel 3: Translation of results; see Supplementary Figure 6), we present several options:

- How much time in different types of activity is necessary for the individual or cohort to reach the 50^th^ percentile or in case they are already above the 50^th^ percentile, how much time is necessary to increase their physical activity by 5%.
- How much average acceleration and intensity gradient need to be increased to correspond to a clinically relevant increase in cardiorespiratory fitness.
- How much average acceleration and intensity gradient need to be increased to correspond to a relevant reduction in mortality and cardiovascular disease risk.

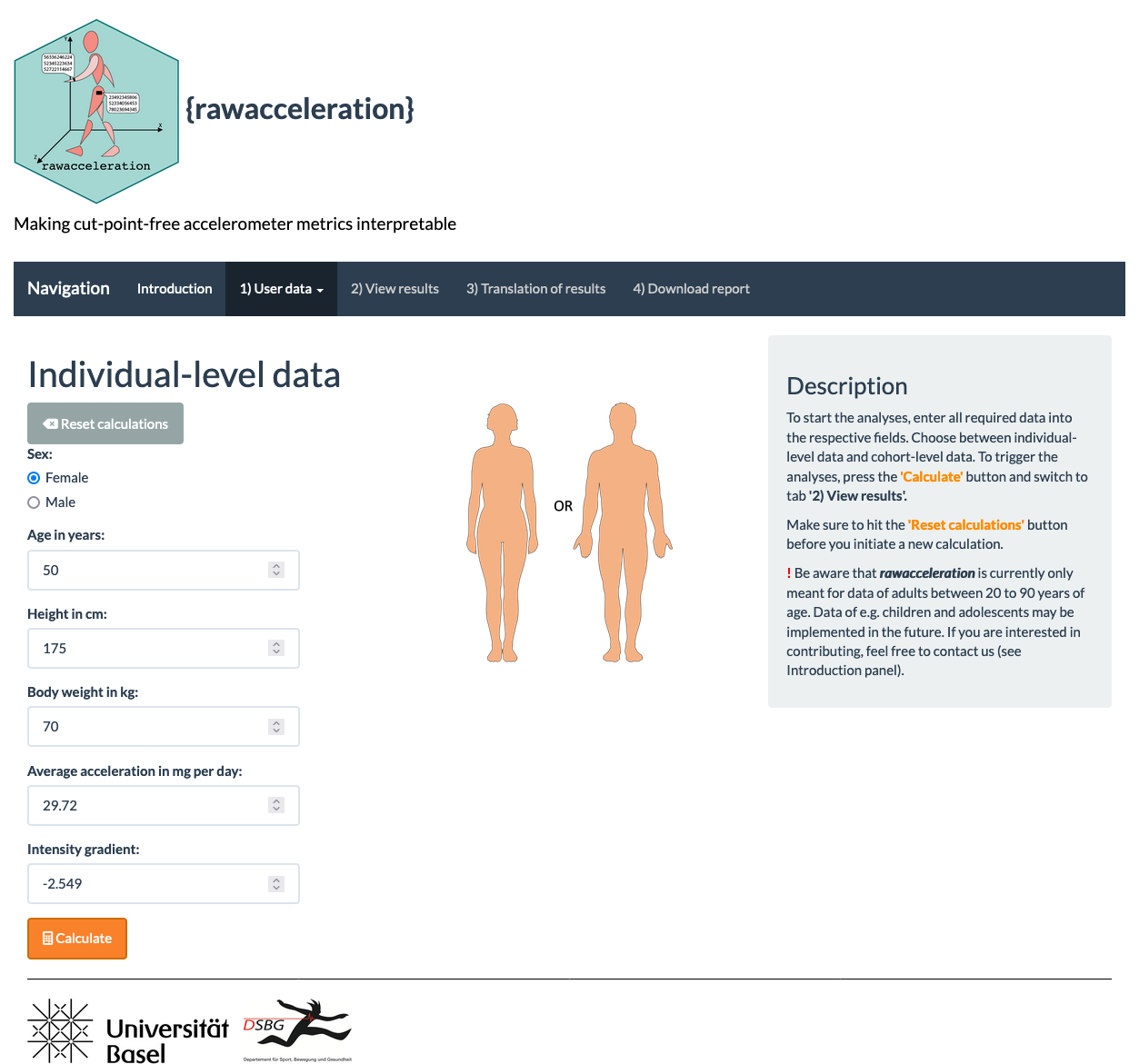

**Supplementary Figure 4.** User input panel. One of three available options (individual-level data, aggregated cohort-level data, or raw data).

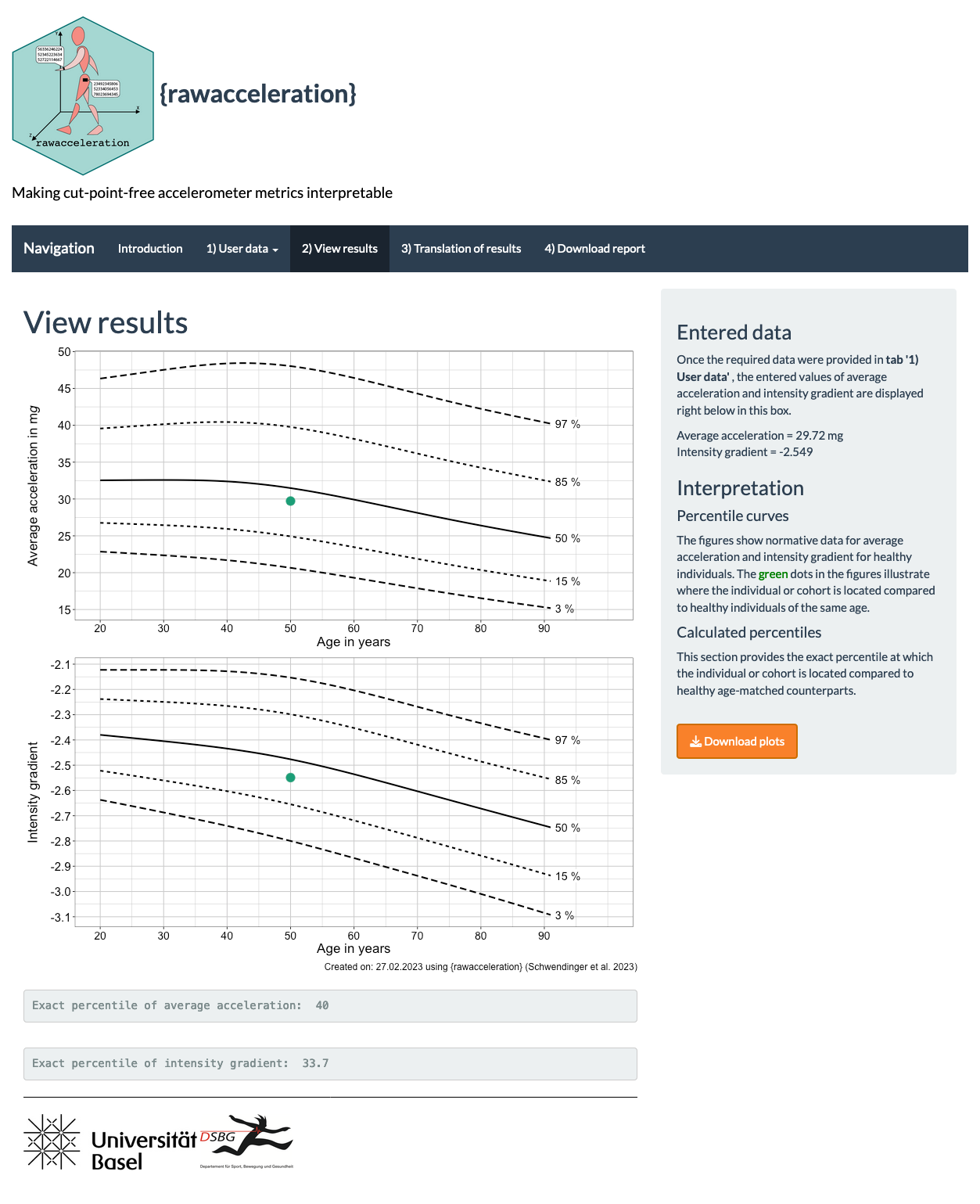

**Supplementary Figure 5.** Results panel. Presents the user data plotted into our percentile curves and yield the exact percentile the individual of cohort is on.

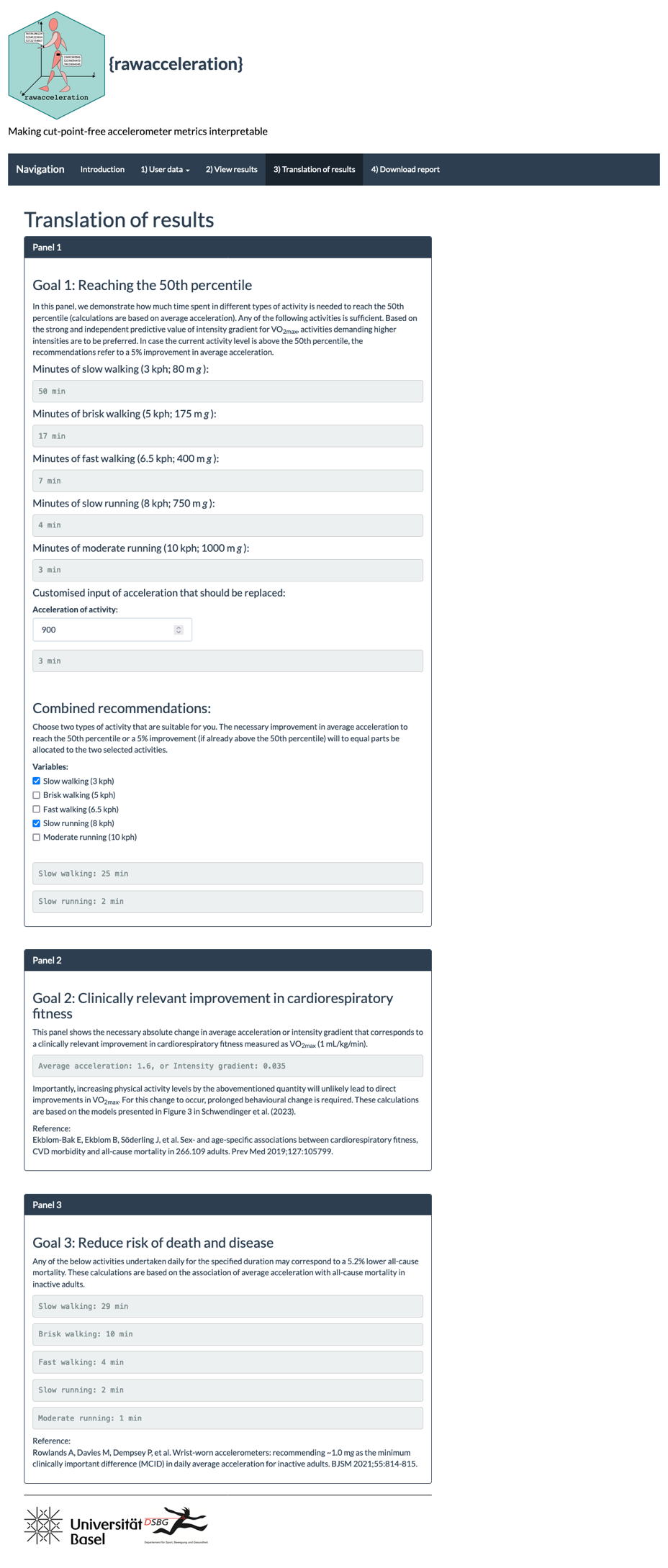

**Supplementary Figure 6.** Translation of results panel. Provides information on how much time in the respective activities is needed to reach the 50^th^ percentile or a 5% increase in activity for those that are already above the 50^th^ percentile.

**References**

1.	Migueles JH, Rowlands AV, Huber F, Sabia S, van Hees VT. GGIR: A Research Community–Driven Open Source R Package for Generating Physical Activity and Sleep Outcomes From Multi-Day Raw Accelerometer Data. Journal for the Measurement of Physical Behaviour. J Meas Phys Behav. 2019;2(3):188-96.

2.	van Hees VT, Gorzelniak L, Dean León EC, Eder M, Pias M, Taherian S, et al. Separating Movement and Gravity Components in an Acceleration Signal and Implications for the Assessment of Human Daily Physical Activity. PloS ONE. 2013;8(4):e61691.

3.	R Core Team. R: A language and environment for statistical computing Vienna, Austria: R Foundation for Statistical Computing; 2021 [Available from: https://www.R-project.org/].

4.	van Hees VT, Fang Z, Langford J, Assah F, Mohammad A, da Silva IC, et al. Autocalibration of accelerometer data for free-living physical activity assessment using local gravity and temperature: an evaluation on four continents. J Appl Physiol. 2014;117(7):738-44.

5.	van Hees VT, Sabia S, Jones SE, Wood AR, Anderson KN, Kivimäki M, et al. Estimating sleep parameters using an accelerometer without sleep diary. Sci Rep. 2018;8(1):12975.

6.	Hildebrand M, van Hees VT, Hansen BH, Ekelund U. Age Group Comparability of Raw Accelerometer Output from Wrist- and Hip-Worn Monitors. Med Sci Sports Exerc. 2014;46(9):1816-24.

7.	Rowlands AV, Mirkes EM, Yates TOM, Clemes S, Davies M, Khunti K, et al. Accelerometer-assessed Physical Activity in Epidemiology: Are Monitors Equivalent? Med Sci Sports Exerc. 2018;50(2):257-265.

8.	Rowlands AV, Edwardson CL, Davies MJ, Khunti K, Harrington DM, Yates T. Beyond Cut Points: Accelerometer Metrics that Capture the Physical Activity Profile. Med Sci Sports Exerc. 2018;50(6):1323-32.

9.	Fairclough SJ, Taylor S, Rowlands AV, Boddy LM, Noonan RJ. Average acceleration and intensity gradient of primary school children and associations with indicators of health and well-being. J Sports Sci. 2019;37(18):2159-67.
